# Safety and anti-inflammatory effects of ILB-202, an engineered extracellular vesicles for NF-κB inhibition: A double-blind, randomized, placebo-controlled phase 1 trial

**DOI:** 10.1101/2025.03.25.25324589

**Authors:** Seoyeon Hyun, Hojun Choi, Yujin Sub, Dasom Hong, So-Hee Ahn, Kyungsun Choi, Seungwook Ryu, Cheolhyoung Park, Heon Yung Gee, Chulhee Choi

**Author notes:** Corresponding authors. Heon Yung Gee, MD, PhD; Department of Pharmacology, Brain Korea 21 PLUS Project for Medical Sciences, Yonsei University College of Medicine, Seoul 03722, Republic of Korea; and Chulhee Choi, MD, PhD; ILIAS Biologics Inc., Daejeon 34014, Republic of Korea. These authors contributed equally: Seoyeon Hyun, Hojun Choi, Yujin Sub.

## Abstract

Excessive activation of NF-κB is implicated in the pathogenesis of numerous inflammatory and autoimmune disease; however, conventional NF-κB inhibitors often cause widespread immunosuppression. In contrast, extracellular vesicles (EVs) are promising vehicles for therapeutic cargo delivery with advantages including reduced risk of replication. In this single-center, randomized, double-blind, placebo-controlled phase 1 trial, we evaluated ILB-202, an engineered, allogeneic EV derived from HEK293 cells and loaded with a super-repressor IκBα. A single ascending intravenous dose of ILB-202 was administered to 18 healthy volunteers, and the safety, tolerability, and preliminary pharmacodynamic effects were assessed. ILB-202 was well tolerated at all dose levels with no serious or dose-limiting toxicities; only minor adverse events, including a mild decrease in NK cell counts and one case of grade 1 neutropenia, were observed. The laboratory parameters, vital signs, and cytokine profiles remained stable, indicating no systemic immunogenicity. Single-cell RNA sequencing revealed subtle, time-dependent modulation of NF-κB-associated pathways, enhanced TGFβ and visfatin signaling and reduced TNF signaling—suggesting a shift toward an anti-inflammatory state. These findings support the safety and immunomodulatory activity of ILB-202 and pave the way for future trials in diseases characterized by dysregulated NF-κB activation. The trial is registered at ClinicalTrials.gov (ID number NCT05843799).

## Introduction

Extracellular vesicles (EVs) are small, membrane-bound structures secreted by virtually all cell types. They carry a diverse spectrum of biomolecular cargo, including lipids, nucleic acids, and proteins, which can modulate the phenotype of target cells (*1*). EV-based therapeutics aim to exploit these natural intercellular communication vesicles to deliver functional cargo to the target cells (*2, 3*). Compared to cell therapies, unmodified (“naïve”) EVs offer a cell-free approach that mitigates risks of replication, tumorigenesis, and ethical challenges (*4*). Moreover, EVs can be engineered to carry active pharmaceutical ingredients (APIs), enhancing intracellular delivery while protecting APIs from enzymatic degradation and antibody-mediated clearance (*5, 6*). To date, the most clinical trials involving EVs have used naïve EVs derived from mesenchymal stem cells, focusing on indications such as tissue repair and immune modulation (*4, 7–11*). However, the clinical development of engineered EV therapeutics remains in its early stages. Three phase 1 clinical trials have explored engineered EVs: ExoSTING (EVs loaded with STING agonists; ClinicalTrials.gov identifier: NCT04592484), exoIL12 (EVs displaying IL-12 on their surface; ClinicalTrials.gov identifier: NCT05156229), and EXO-CD24 (EVs displaying CD24) (*12*). Although these studies highlight the potential of engineered EVs, their intratumoral or inhalation delivery routes limit the assessment of their systemic immunogenicity and safety profiles.

ILB-202, an EV designed to inhibit NF-κB, is engineered to carry super-repressor IκBα (srIκB), a stabilized version of the NF-κB inhibitor protein IκBα with an extended half-life (*13, 14*). NF-κB is a pivotal transcription factor that governs both innate and adaptive immune responses (*15*). Excessive activation of this pathway had been implicated in the pathogenesis of numerous inflammatory and autoimmune diseases, making it a prime therapeutic target (*16*). However, conventional NF-κB inhibitors suffer from specificity issues and often cause widespread immunosuppression due to their broad blockade of basal NF-κB signaling, including those functioning within normal physiological ranges (*17–19*). In contrast, ILB-202 was designed to suppress NF-κB primarily in cells exhibiting pathologically high NF-κB activity. Under normal conditions, endogenous IκBα restrains NF-κB, thereby preventing unchecked inflammation. However, when pro-inflammatory signals arise, IκBα is rapidly phosphorylated and degraded, freeing NF-κB to translocate to the nucleus and activate pro-inflammatory genes. The srIκB cargo in ILB-202 is resistant to this degradation, selectively counteracting hyperactive NF-κB in inflamed tissues while having minimal effect on healthy, non-inflamed cells (*14, 20*). Preclinical studies have demonstrated that ILB-202 effectively mitigates inflammation across various disease models, including sepsis, kidney ischemia-reperfusion injury, chronic post-ischemia pain, rheumatoid arthritis, inflammation-induced preterm birth, and age-related neuroinflammation (*20–25*).

Therefore, in this single-center, randomized, double-blind, placebo-controlled phase 1 clinical trial, we evaluated the safety and tolerability of ILB-202 administered as a single ascending dose via intravenous infusion in healthy volunteers, focusing on its immunogenicity, dose-limiting toxicities, and preliminary pharmacodynamic effects. Additionally, we evaluated potential biomarkers associated with the therapeutic efficacy of ILB-202, offering insights into its mechanism of action at the cellular and molecular levels. To the best of our knowledge, this is the first clinical trial investigating systemically administered allogenic EVs.

## 2. Materials and Methods

### 2.1 Study design

This phase I, randomized, double-blind, placebo-controlled, first-time-in-human single ascending dose study (ILIAS study ILB-202-001; NCT05843799) was conducted at a single center in Australia between May 09, 2023, and Oct 04, 2023. We enrolled healthy participants into three ILB-202 dose cohorts: Cohorts 1–3.

The participants were randomized into treatment groups with sentinel dosing (one active dose and one placebo for each cohort). ILB-202 (or placebo) was administered via a 2 intravenous (IV) infusion (infusion rate of 250 mL/120 min). The required volume of ILB-202 (5 × 10¹¹ particle number [pn]/mL) was diluted with 0.9% sodium chloride in a 250 mL infusion bag for each dose level. The volume of ILB-202 (determined based on the assigned dose level and the participant’s weight measured on day −1) was aseptically added to the IV infusion bag after removing an equivalent volume of saline to maintain blinding.

ILB-202, was prepared by aseptically adding 4 mL of ILB-202 vehicle to an infusion bag containing 250 mL of normal saline. Before adding ILB-202, an equivalent volume of saline was removed to maintain consistency. Participants assigned to receive the placebo received a single IV infusion bag containing 250 mL of normal saline.

### 2.2 Preparation of clinical grade of ILB-202

The production of ILB-202 employs a light-induced heterodimerization system using genetically engineered HEK293 cells. These cells are transfected with a pcDNA3.1(+) plasmid vector encoding srIκB-CRY2 and CIBN-CD9. Under blue light illumination, CRY2 transiently binds to CIBN, a truncated form of CIB1 fused to EV-associated tetraspanin CD9. This interaction directs the cargo proteins to the surface of the plasma membrane of the early endosomes, facilitating the encapsulation of srIκB into the EVs. The resulting multivesicular bodies fuse with the plasma membrane, releasing EVs loaded with the CRY2-conjugated cargo (*26*).

The manufacturing process began with cell culture using the HEK293 cell line derived from at Master Cell Bank. Characterization and testing of the Master Cell Bank were conducted by Sartorius (1 Technology Terrace, Todd Campus, West Scotland Science Park, Glasgow, UK) in accordance with the ICH Q5D guidelines.

ILB-202 drug substance (DS) was manufactured under Good Manufacturing Practice (GMP) conditions at KBIO Health (Cheongju-si, Republic of Korea). Cell culture was carried out in four-wave bioreactors, purchased from Sartorius, under blue light illumination with a total working volume of approximately 100 L for 4 days. At the end of the production cycle, the cells and cellular debris were removed by centrifugation at 2,000 g for 10 min. The harvested material was further purified by ultrafiltration and diafiltration to concentrate the solution, perform buffer exchange, and facilitate anionic and multi-modal resin chromatography. Finally, sterile filtration was performed to reduce bioburden before the drug substance was frozen and stored at −70°C.

The ILB-202 drug product (DP) was also manufactured under GMP conditions; however, this step was performed at the Korea Bio Pharmaceutical CMO Center (Andong-si, Republic of Korea). The frozen drug was transferred from KBIO Health to the manufacturing site where it was thawed and prepared for the final DP formulation. After formulation, the product was subjected to sterile filtration before being filled into vials at a nominal fill volume. The finished DP was then stored at −70°C until it was ready for distribution to support clinical trials.

General properties such as appearance and pH, particle and protein concentration of ILB-202, purity, and drug potency were assessed to evaluate the quality of the ILB-202 DS and DP. Additionally, endotoxin, adventitious virus, sterility, and bioburden tests were performed to confirm safety. All tests confirmed that the ILB-202 DS and DP meet the specified acceptance criteria, ensuring that no quality issues are present and that the product was safe for use in clinical trials.

### 2.3 Study population

Only healthy volunteers who met the specified inclusion and exclusion criteria participated in this study (**Table 1**, the complete inclusion and exclusion criteria are provided in **Supplementary Table 1**). Participants eligible for the study were healthy, 18–55 years of age, with a body mass index (BMI) ≥18.0 ≤30.0 kg/m^2^ at screening. Participants were excluded if they were unable to refrain from, or anticipated the use of, any prescription medications or were administered over-the-counter drugs, dietary supplements, or herbal remedies within 14 days or five half-lives prior to dosing and throughout the study.

**Table 1.**
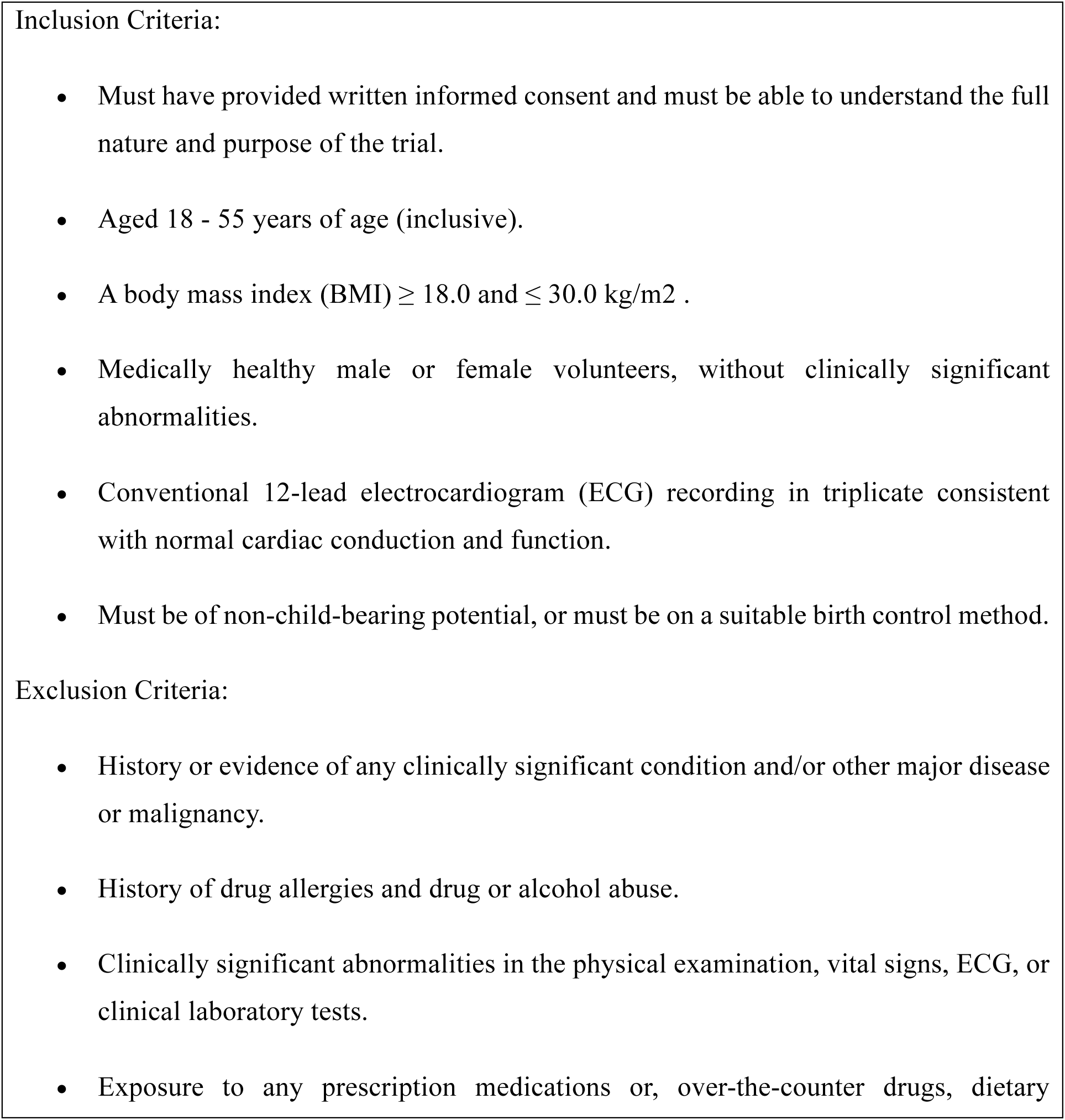
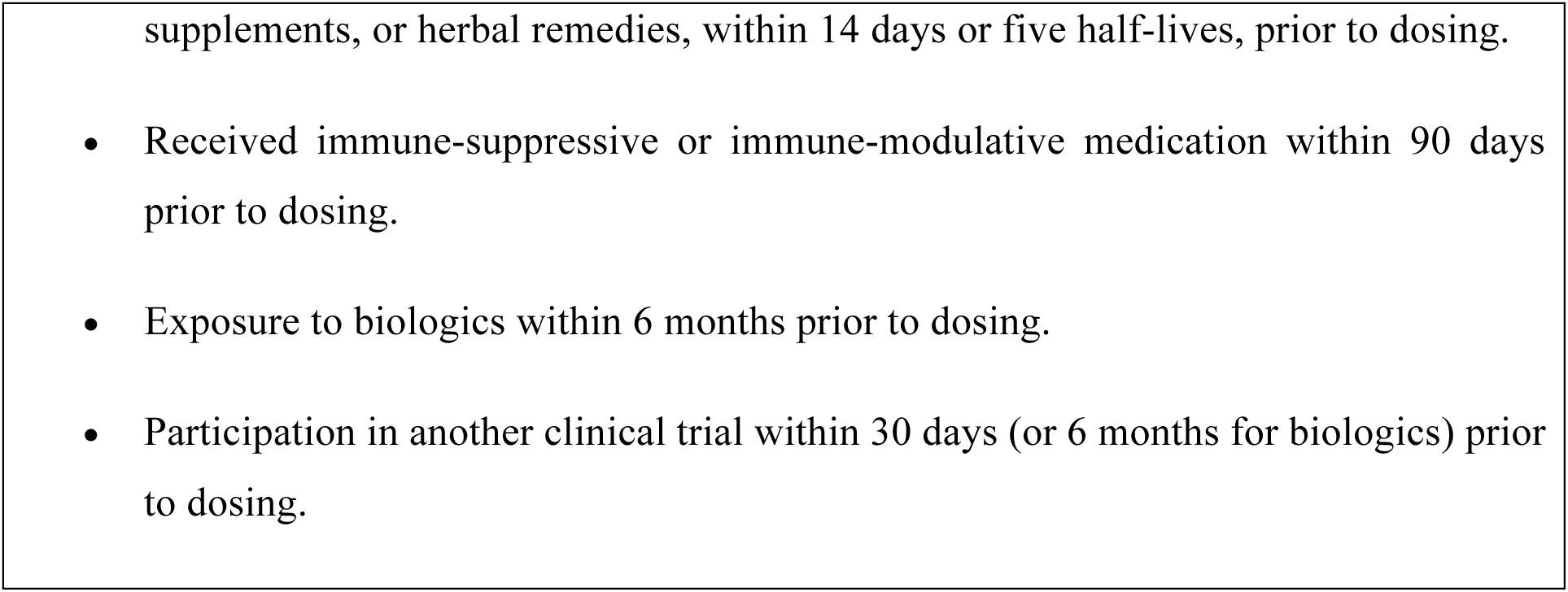
Summary of inclusion and exclusion criteria.

Participants were also excluded if they had received treatment with immunosuppressive or immunomodulatory medication (including topical, systemic, and inhalant corticosteroids) or had received immunoglobulins and/or blood products within 90 days and biologics within 6 months prior to the first dosing and throughout the study (for further details, see **Supplementary Table 2**). Participants who took a prohibited medications during the study were required to notify the principal investigator promptly, and details of medication use were documented. If the use of any medication was deemed necessary by the PI, a discussion with the sponsor was conducted.

### 2.4 Safety and tolerability

The primary objective of the study was to assess the safety and tolerability of ILB-202 by monitoring adverse events (AEs), vital signs, body weight, physical examinations results, routine hematology, clinical chemistry, urinalysis, and electrocardiography (EGG). Serum cytokine levels (IL-1β, IL-2, IL-6, IL-10, IFN-γ, and TNF-α) and C-reactive protein (CRP) were assessed for 24 h. Peripheral blood CD4+ (T_helper_), CD8+ (T_cytotoxic_), CD19+ (B-cells), and CD16/56+ (natural killer [NK] cells) were measured using flow cytometry to assess markers of immune cell activation following a single IV infusion of ILB-202. Another exploratory objective of the study was to collect and retain samples of plasma, red blood cell (RBC)-lysed whole blood, and peripheral blood mononuclear cells (PBMCs) samples for potential future evaluation of the pharmacokinetics and pharmacodynamics of ILB-202.

### 2.5 Blood sampling

Blood samples were collected at predefined time points, pre- and post-study drug administration to evaluate the pharmacodynamic properties and pharmacokinetics of ILB-202.

### 2.6 Single-cell RNA sequencing (scRNA-seq) analysis

#### 2.6.1 Single-cell preparation

Whole peripheral venous blood was collected in K_2_EDTA Vacutainer tubes. PBMCs were isolated from the collected blood using Ficoll-Paque density gradient centrifugation with V-bottom tubes. The isolated PBMCs were cryopreserved in a freezing medium at a density of 1–4 × 10^7^ cells/mL. The PBMCs were placed in Mr Frosty containers and transferred to Ilias Biologics (Daejeon, South Korea)/Rokit Genomics (Seoul, South Korea) at nominal −80 ℃ and subsequently stored in liquid nitrogen. Cell stocks were thawed in 37℃ 10% FBS/DMEM. Cells were washed twice with cold Ca^2+^ and Mg^2+^ free 0.04 % BSA/PBS at 300g for 5 min at 4°C. Samples were gently resuspended in cold Ca^2+^ and Mg^2+^ free 0.04% BSA/PBS, and stained with acridine orange and propidium. The cells were counted using a LUNA-FX7™ Automated Fluorescence Cell Counter (Logos biosystems, Anyang, South Korea.

#### 2.6.2 Multiplexing individual samples for scRNA-seq

Each sample was tagged with an antibody-polyadenylated DNA barcode for the human cells. Briefly, the cells were stained with a multiplexing antibody for 20 min at room temperature and washed three times by using with staining buffer. After the final wash, the samples were gently resuspended in a cold sample buffer, counted using a LUNA-FX Automated Fluorescence Cell Counter (Logos Biosystems), and pooled.

#### 2.6.3 scRNA-seq library construction

Single-cell capture was performed using BD Rhapsody Express instrument according to the manufacturer’s instructions (BD Bioscience). Briefly, cells pooled from each sample in cold sample buffer were loaded into the BD Rhapsody cartridge. After cell separation, the cell-barcode magnetic beads were loaded into the cartridge. Cells were lysed and mRNA capture beads were retrieved. cDNA synthesis and Exonuclease I treatment were performed on the mRNA capture beads using the BD Rhapsody cDNA kit.

#### 2.6.4 Library preparation for scRNA-seq

scRNA-seq libraries were constructed using the BD Rhapsody whole-transcriptome analysis (WTA) amplification kit according to the ‘mRNA Whole Transcriptome Analysis and Sample Tag Library Preparation’ protocol. Briefly, cDNA was sequentially subjected to random priming and extension (RPE), RPE amplification, and index PCR for WTA library contruction. Additionally, cDNA was sequentially subjected to nested PCR (PCR 1 and 2) and an index PCR for the sample tag library.

#### 2.6.5 scRNA-seq

Purified WTA and sample tag libraries were quantified using qPCR according to the qPCR Quantification Protocol Guide (KAPA) and qualified using an Agilent Technologies 4200 TapeStation (Agilent technologies). Libraries were then pooled and sequenced using the HiSeq platform (Illumina) to generate 150 bp paired-end reads. The sequencing depth of the WTA library was approximately 20,000 reads/cell, and that of the sample tag library was approximately 400 reads/cell.

#### 2.6.6 Data analysis

scRNA-seq outputs were processed and mapped to the human reference genome (GRCh38) using the proprietary pipeline from ROKIT Genomics (Seoul, Korea). The resulting data were produced in matrix format, containing gene expression counts for downstream analysis. Gene-wise read counts for genes with a minimum of 1,500 reads were exported from the ROKIT Genomics pipeline in Matrix Market format and subsequently imported into R using a custom script compatible with Seurat’s data input functions (v5.1.0). An average of >34 million reads from > 5,800 cells were obtained for each sample. Samples were evaluated for quality, and the cells were filtered based on the following commonly used criteria to ensure high-quality cells, including thresholds for gene counts, unique molecular identifier (UMI), and the proportion of mitochondrial gene expression: cohort1, gene count >250, UMI >2,000, and mitochondrial gene expression <20%; cohort2, gene count >600, UMI >4500, and mitochondrial gene expression <20%; cohort3, gene count >300, UMI >1,500, and mitochondrial gene expression <25% (*27*). Subseqeuntly, 122,956 cells were retained: 29,133, 1, 17,718, and 76,105 cells from cohort 1, 2, and 3, respectively. The samples were combined using an algorithm implemented in Seurat using the default function settings with principal component dimensions of 1:20 for all dimension reduction and integration steps. Batch effects across cohorts 1, 2, and 3 were corrected using the harmony algorithm implemented in Seurat. Principal component analysis (PCA) embeddings were adjusted to remove batch-specific variations while preserving biological signals. The corrected data were subsequently used for downstream analyses, including clustering and differentially expressed genes (DEGs). The cluster resolutions were set to 0.5 unless otherwise stated. The RunUMAP random seed was set to 1,000 to ensure reproducibility. Marker genes for cell clusters were identified using the Seurat FindmArkers function with default parameters. Cell type annotation was performed using the SingleR package (v2.6.0) with the HumanProteinAtlas database from the Celldex package (v1.14.0). However, some NK and T cells were inconsistently annotated based on marker gene expression. Consequently, re-annotation was performed using the expression values of CD3 (T cell marker) and NCAM1 (NK cell marker) to ensure more accurate classification. DEGs analysis was initially performed using stringent criteria, specifically an FDR corrected p < 0.001 and |log_2_(fold change)| ≥ 1, across three-time points: pre-injection (Pre) versus at the end-of-infusion (EOI), EOI versus 24 h after infusion (24h), and Pre versus 24h. Consequently, owing to the restricted number of DEGs identified under these criteria, a more lenient threshold was utilized for KEGG pathway analysis. Genes with a raw p <0.05 and |log_2_(fold change)| >0 were utilized for KEGG pathway enrichment analysis to provide comprehensive pathway-level insights. Gene set enrichment analysis (GSEA) and KEGG pathway analysis were conducted using the gseGO and enrichKEGG functions from clusterProfiler (v4.10.1), respectively. GSEA plots were generated from the DEG data using the gseaplot function. Gene set scores of the monocyte subtype markers were calculated for each cell type using the AddModuleScore function in Seurat. Cell-cell interaction analysis was performed using the CellChat package (v1.6.1) to identify significant intercellular communications at each time point (Pre, EOI, and 24h). CellChat was run independently for each time point, and significant interactions were extracted using the subset communication function. This allowed the identification of cell-cell interactions that were specific to a particular time point but were not deemed significant at other times. Interactions were further categorized into source-, and target-specific cell types to emphasize time point-specific patterns. The results were visualized using netVisual_bubble and netVisual_aggregate plots, offering an intuitive representation of distinct and significant interactions at each time point.

### 2.7 Ethics

All participants provided written informed consent after the aims and methods of the study were explained according to their level of knowledge. This study was reviewed and approved by an appropriate human research ethics committee before the study was initiated. The requirements for conducting clinical trials in accordance with the applicable regulations of the Australian Therapeutic Goods Administration under the Clinical Trial Notification scheme were met before commencement of this study.

This study was conducted according to the principles of the Declaration of Helsinki, International Council for Harmonization of Technical Requirements for Pharmaceuticals for Human Use Guideline for Good Clinical Practice E6(R2) (2016) (as adopted in Australia) and the Australian National Health and Medical Research Council National Statement on Ethical Conduct in Human Research (2007, incorporating all updates). The current investigation was registered at ClinicalTrials.gov (ID number NCT05843799).

### 2.8 Sample size and statistical analysis

As the primary endpoints of the study was safety and tolerability, the sample size was based on feasibility, and no formal sample size calculation was performed for this first-time-in-human study with ILB-202. Six subjects per cohort were considered sufficient to describe the safety and tolerability of ILB-202. Thus, three planned sample size was a total of 18 participants.

## 3. Results

### 3.1 Participant disposition and demographics

Eighteen healthy participants (n = 37) were randomized into the three cohorts and completed the study (**Fig. 1**). The baseline patient characteristics are summarized in **Table 2**.

**Figure 1.**
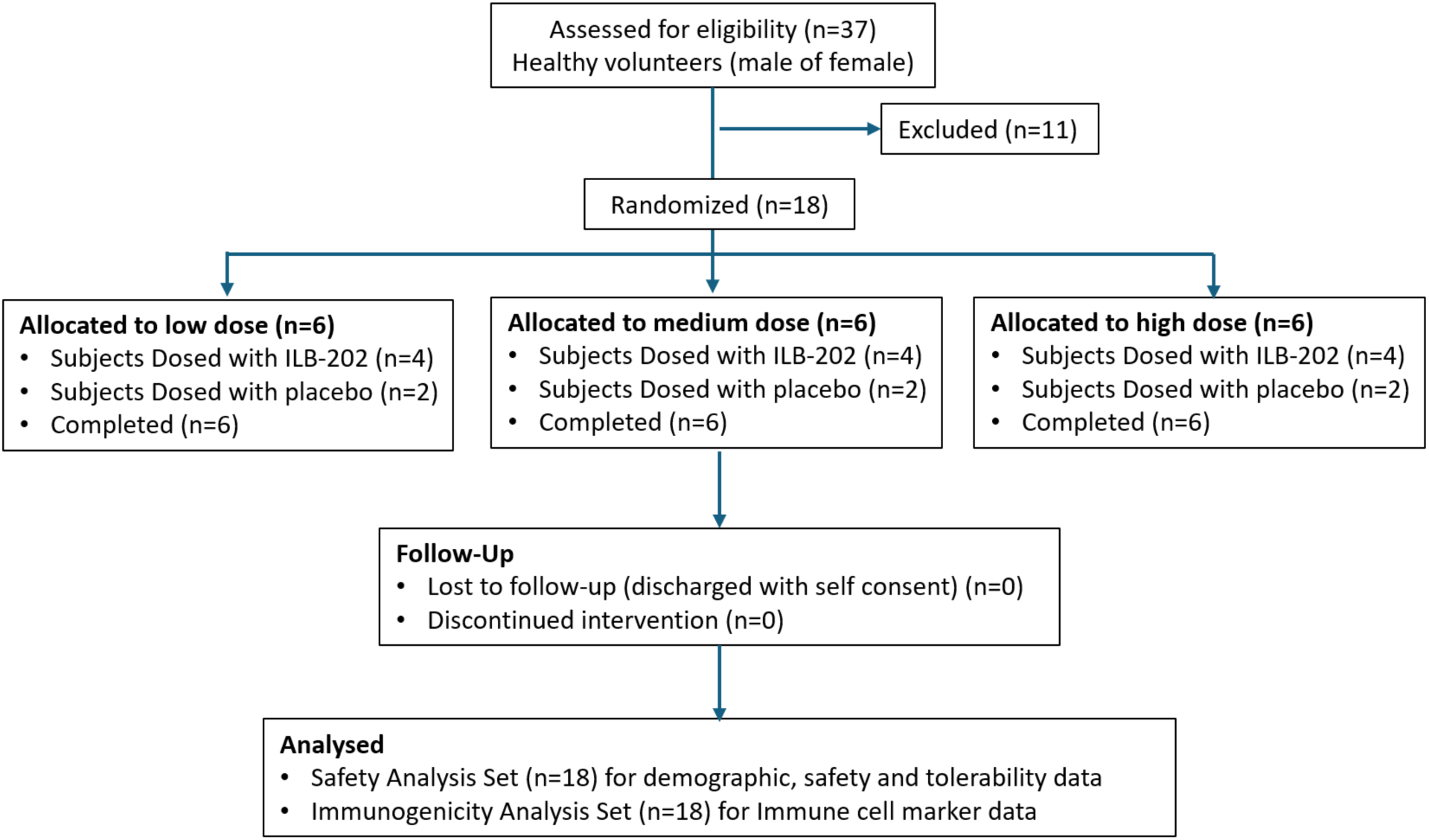
Participant enrollment and study flow diagram.

This flow diagram outlines the enrollment, randomization, dosing, and analysis of participants in the phase 1 clinical trial of ILB-202. Of the 37 individuals assessed for eligibility, 18 were randomized into three dose groups: low dose (n=6), medium dose (n=6), and high dose (n=6). Each group included four participants receiving ILB-202 and two who received placebo. All randomized participants completed the study, with no cases of loss to follow-up or discontinuation of the intervention. Safety, tolerability, and immune cell marker data were analyzed in all 18 participants.

**Table 2.** Summary of demographics.

| Demographic Parameter | Statistic | Sub-group | Pooled Placebo (n=6) | ILB-202 Low dose (n=4) | ILB-202 Medium dose (n=4) | LB-202 High dose (n=4) | Pooled ILB-202 (n=12) | Overall (n=18) |
| --- | --- | --- | --- | --- | --- | --- | --- | --- |
| Age (years) | Mean (± SD) | - | 23.0 (2.19) | 34.8 (3.59) | 31.5 (13.13) | 36.5 (14.62) | 34.3 (10.65) | 30.5 (10.23) |
|  | Mean | Female | 23 | 34 | 29 | 53 | 38.7 | 28.9 |
| Sex | n (%) | Male | 1 (16.7) | 3 (75.0) | 3 (75.0) | 3 (75.0) | 9 (75.0) | 10 (55.6) |
|  |  | Female | 5 (83.3) | 1 (25.0) | 1 (25.0) | 1 (25.0) | 3 (25.0) | 8 (44.4) |
| Woman of child-bearing potential | n (%) | Yes | 5 (100.0) | 1 (100.0) | 1 (100.0) | - | 2 (66.7) | 7 (87.5) |
|  |  | No | - | - | - | 1 (100.0) | 1 (33.3) | 1 (12.5) |
| Height (cm) | Mean (± SD) | - | 175.90 (9.560) | 171.50 (7.400) | 172.20 (6.600) | 170.30 (6.920) | 171.30 (6.370) | 172.90 (7.620) |
| Weight (kg) | Mean (± SD) | - | 77.30 (11.560) | 74.50 (17.960) | 73.10 (13.320) | 80.10 (6.360) | 75.90 (12.550) | 76.40 (11.900) |
| BMI (kg/m<sup>2</sup>) | Mean (± SD) | - | 24.90 (2.860) | 25.10 (4.290) | 24.50 (3.580) | 27.60 (1.520) | 25.70 (3.330) | 25.50 (3.120) |
| Race | n (%) | White | 6 (100.0) | 3 (75.0) | 1 (25.0) | 2 (50.0) | 6 (50.0) | 12 (66.7) |
|  |  | Asian | - | 1 (25.0) | 1 (25.0) | 1 (25.0) | 3 (25.0) | 3 (16.7) |
|  |  | Black or African American | - | - | 1 (25.0) | 1 (25.0) | 2 (16.7) | 2 (11.1) |
|  |  | Multiple | - | - | 1 (25.0) | - | 1 (8.3) | 1 (5.6) |
| Ethnicity | n (%) | Hispanic or Latino | - | 1 (25.0) | - | - | 1 (8.3) | 1 (5.6) |
|  |  | Not Hispanic or Latino | 6 (100.0) | 3 (75.0) | 4 (100.0) | 4 (100.0) | 11 (91.7) | 17 (94.4) |

Abbreviations: BMI = Body mass index; SD = Standard deviation. Notes: Denominator for participant of child-bearing potential was based on the count of female participants.

### 3.2 Safety and tolerability

No treatment-emergent AEs exceeding grade 1 were noted after a single IV dose of ILB-202, including high dose. Additionally, no TEAEs resulted in mortality or were serious, no participant withdrew from the study or study drug because of AEs. AEs were reported in 10 of 12 (83.3%) healthy participants receiving ILB-202 and two of six (33.3%) receiving a placebo (**Table 3 and Supplementary Table 3**). No clinically significant pattern was observed in vital signs, body weight, ECGs, or laboratory parameters, including serum IL-1β, IL-2, IL-6, IL-10, IFN-γ, and TNF-α, or CRP.

**Table 3.** Summary of treatment-emergent adverse events (Safety analysis set)

|  | **Pooled**<br>**Placebo**<br>**(n=6)**<br>**n (%)** | **ILB-202**<br>**Low dose**<br>**(n=4)**<br>**n (%)** | **ILB-202**<br>**Medium**<br>**dose**<br>**(n=4)**<br>**n (%)** | **ILB-202**<br>**High dose**<br>**(n=4)**<br>**n (%)** | **Pooled**<br>**ILB-202**<br>**(n=12)**<br>**n (%)** | **Overall**<br>**(n=18)**<br>**n (%)** |
| --- | --- | --- | --- | --- | --- | --- |
| **Overall** |  |  |  |  |  |  |
| Number of participants with: |  |  |  |  |  |  |
| TEAE ≥ 1 | 2 (33.3) | 3 (75.0) | 3 (75.0) | 4 (100.0) | 10 (83.3) | 12 (66.7) |
| Serious TEAE ≥ 1 | - | - | - | - | - | - |
| Serious TEAE leading to study discontinuation ≥ 1 | - | - | - | - | - | - |
| TEAE leading to study drug discontinuation ≥ 1 | - | - | - | - | - | - |
| TEAE leading to death ≥ 1 | - | - | - | - | - | - |
| **TEAEs by relationship to study drug** |  |  |  |  |  |  |
| Number of participants with at least 1 TEAE: |  |  |  |  |  |  |
| Not related | 1 (16.7) | - | - | 1 (25.0) | 1 (8.3) | 2 (11.1) |
| Unlikely related | - | 3 (75.0) | - | 2 (50.0) | 5 (41.7) | 5 (27.8) |
| Possibly related | 1 (16.7) | - | 3 (75.0) | 2 (50.0) | 5 (41.7) | 6 (33.3) |
| Probably related | - | - | - | - | - | - |
| Definitely related | - | - | - | - | - | - |
| **TEAEs by severity** |  |  |  |  |  |  |
| Number of participants with at least 1 TEAE: |  |  |  |  |  |  |
| Mild (Grade 1) | 1 (16.7) | 3 (75.0) | 3 (75.0) | 4 (100.0) | 10 (83.3) | 11 (61.1) |
| Moderate (Grade 2) | 1 (16.7) | - | - | - | - | 1 (5.6) |
| Severe (Grade 3) | - | - | - | - | - | - |
| Life-threatening (Grade 4) | - | - | - | - | - | - |
| Death (Grade 5) | - | - | - | - | - | - |

Abbreviations: TEAE = treatment-emergent adverse event; n = Number of participants reporting at least one TEAE in that category. Notes: Treatment-emergent AEs are defined as AEs that occurred or worsened from the first administration of study drug up to 8 (+3) days after study drug administration. If a participants experienced the same AE multiple times, this was only to be counted once per System Organ Class and Preferred Term to count the number of participants experiencing that AE. Adverse events that were experienced multiple times with different relationships/severities were only to be counted once per System Organ Class and Preferred Term for the worst-case relationship with study drug/severity.

Abnormalities recorded in the parameters of serum chemistry, urinalysis, vital signs, and ECG were classified by the principal investigator (PI) as non-clinically significant (NCS) and did not meet the criteria of an AE. Hamatological abnormalities were generally deemed NCS; however, one participant in the LB-202 Cohort 3 experienced a low neutrophil count, which was considered clinically significant (CS). This was reported as a TEAE (neutropenia), classified as mild in severity, and deemed possibly related to the study drug. At the time of the last post-study unscheduled follow-up visit, conducted 3 weeks after the end of study visit, this event remained unresolved.

No abnormal coagulation results, obvious changes in inflammatory biomarkers, or changed in body weight over time. Furthermore, no AEs of special interest, such as infusion-related reactions or clinical signs or symptoms indicative of cytokine release syndrome, were reported.

### 3.3 Immunogenicity

No biologically meaningful change was observed in peripheral blood CD4+ (T_helper_), CD8+ (T_cytotoxic_), CD19+ (B-cells), and CD16/56+ (NKcells) levels. However, a decrease was observed from baseline in the percentage of CD16/56+ grandparent cells following administration of ILB-202 at medium and high doses (**Supplementary Table 4**). Despite these minor changes, no indications were observed which suggested that ILB-202 was immunogenic or toxic at the administered doses.

### 3.4 scRNA-seq analysis of PBMCs

Blood samples were collected from two participants who awere administered low and medium doses of ILB-202, while three participants received the high dose, at three-time intervals: Pre, EOI, and 24h. PBMCs were isolated from blood samples, which were then subjected to scRNA-seq to examine their gene expression profiles. Following quality control (QC) filtering (*27*), 122,956 cells were obtained from the blood samples of seven individuals (**Supplementary Fig. 1A-D**). Batch correction and uniform manifold approximation and projection (UMAP) demonstrated well-integrated cells devoid of biases across cohorts, individuals, or time points (**Fig. 2A**). Cell type annotation identified five major cell types: T cells, B cells, NK cells, pre-B cells, monocytes, and hematopoietic stem cells (HSCs); however, neutrophils were absent, likely due to the PBMC isolation method utilized in this study (**Fig. 2A**). The overall populations of these cell types were within the normal range (**Supplementary Fig. 2A**). We monitored the dynamics of PBMC populations for each participant to evaluate whether ILB-202 injection altered cell type proportions. However, no significant increasing or decreasing trends were observed for any particular cell type except for a minor decrease in the NK cell population (**Supplementary Fig. 2B**). Furthermore, DEG analysis was performed to assess significant gene expression alterations between the Pre and EOI time points, as well as between the EOI and 24h time points. No significant differences were identified, irrespective of whether the analysis encompassed the entire cell population or focused on certain cell types (**Fig 2B**, **Supplementary Fig. 2C**). These results indicated that ILB-202 does not significantly affect the physiology of healthy individuals, indicating a potentially desirable safety profile.

**Figure 2.**
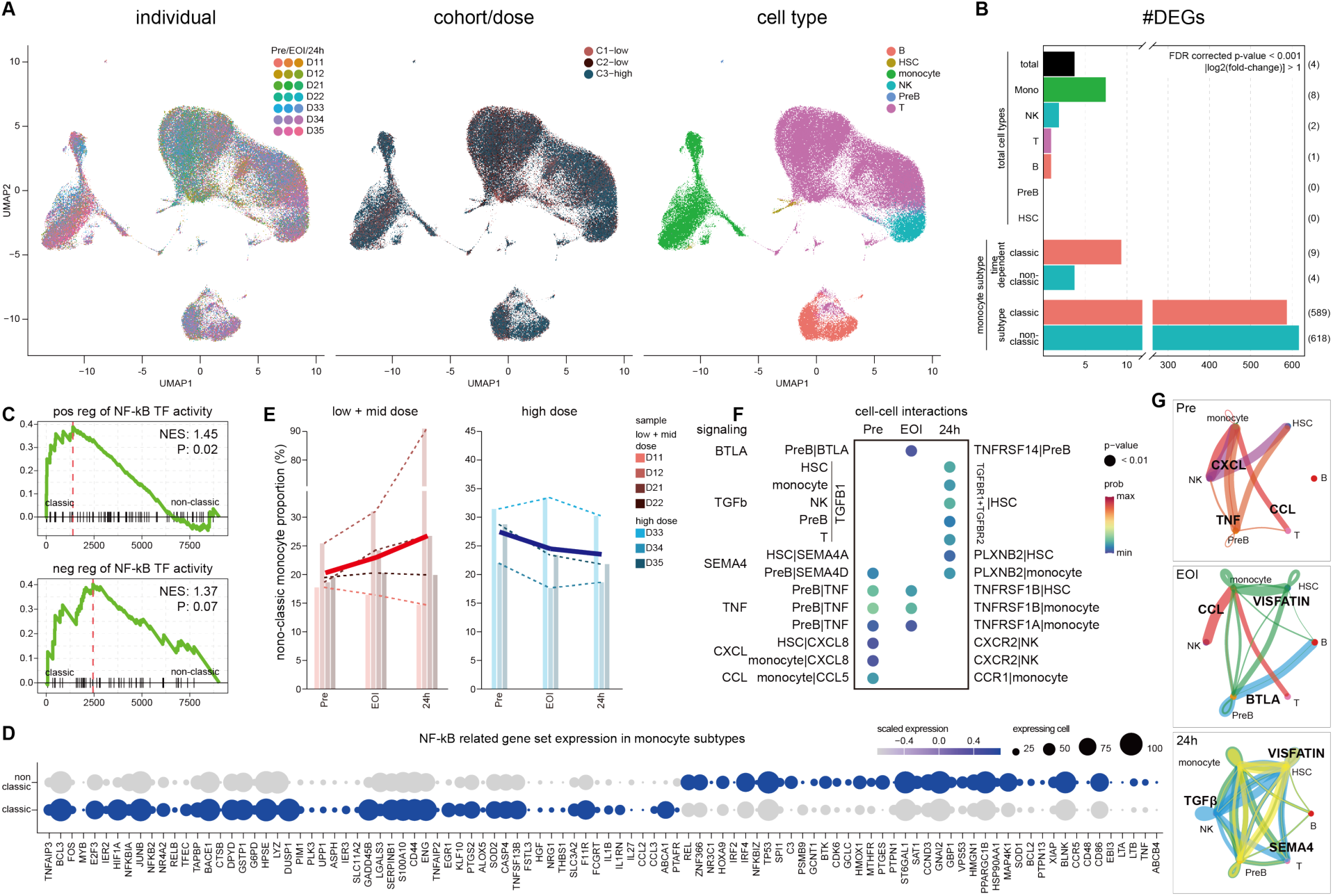
Single-cell transcriptomic analysis of the safety and anti-inflammatory effects of ILB-202 in healthy individuals. **A**. Uniform manifold approximation and projection (UMAP) of scRNA-seq cells (n=122,956 cells) illustrating three different color coding schemes: (1) individual sample time points categorized as pre-injection (Pre), end of infusion (EOI), and 24-h post-injection (24h); (2) experimental cohorts differentiated by drug dosage levels (C1-low, C2-low, C3-high); and (3) annotated cell types, including hematopoietic stem cell (HSC), monocyte, NK, Pre-B, B, and T cells. **B**. Bar plot showing the number of differentially expressed genes (DEGs) identified across the time points. Total represents DEGs without cell type distinction (Pre vs. EOI, EOI vs. 24h), whereas the other bars illustrate DEGs within specific cell types. Time-dependent bars represent DEGs within the classic and non-classic subtypes over time, whereas subtype bars represent DEGs between the classic and non-classic subtypes. DEGs were identified with |log_2_(fold change)| >1 and an adjusted p <0.001. **C**. Gene set enrichment analysis (GSEA) results showing enrichment of genes associated with the positive and negative regulation of NF-κB transcription factor activity, contrasting classic and non-classic monocytes. **D**. Dot plot displaying NF-κB-related genes exhibiting statistically significant differential expression between classic and non-classic monocytes. The sizes of the dots indicate the proportion of expressing cells, whereas the color denotes the scaled expression levels. **E**. Bar plot showing the proportion of non-classic monocytes over time. The bars in red represent the proportions under low- and mid-dose conditions, whereas the blue shades represent high-dose conditions. Superimposed lines show changes in non-classic monocyte proportions for individual samples over time (Pre, EOI, and 24h), highlighting both individual variability in response to dosage and overall temporal progression. Dashed lines indicate individual trajectories; bold solid lines indicate average trends. **F**. Dot plot illustrating the probabilty of cell-cell interaction over time points. Signaling pathways are listed on the left, with source cell types and their receptors on the left side of the plot and target cell types and their ligands on the right. The dot color represents the probability of interaction, whereas the dot size reflects the significe level (p-value). The plot highlights the dynamic signaling interactions among cell types at various time points. **G**. Chord plot showing cell type interactions via signaling pathways specific to each time point. Line thickness signifies the interaction strength, with red representing inflammation-promoting pathways, blue indicating inflammation-resolving pathways and green/yellow denoting neutral pathways. The plot emphasizes time-specific interactions, exemplified by CCL signaling occurring only at the Pre time point.

An analysis employing more lenient criteria was performed to examine the overarching trends in gene expression, which identified an increased number of DEGs (**Supplementary Fig. 2D**). Gene ontology (GO) analysis indicated that the KEGG NF-κB signaling pathway was consistently enriched across multiple cell types, suggesting that this pathway may be widely influenced by ILB-202 (**Supplementary Fig. 3E**).

Among the examined cell types, monocytes exhibited the highest number of DEGs, with eight detected (**Fig. 2B**). Subsequently, we conducted additional detailed analyses of monocytes, focusing on classic and non-classic subtypes. Classic monocytes (CD14^high^/CD16^low^), the predominant subset of circulating monocytes, primarily engage in phagocytic and inflammatory responses (*28*). Non-classic monocytes (CD14^low^/CD16^high^) are associated with tissue repair and immune surveillance (*28*). Monocytes were classified into classic and non-classic subtypes based on the expression levels of CD14 and CD16 (**Supplementary Fig. 3A**). DEG analysis of the two subtypes revealed 589 and 618 DEGs in classic and non-classic monocytes, respectively (**Fig. 2B**). GSEA of these subtype-specific DEGs revealed significant enrichment in NF-κB-related activity, indicating functional divergence between the two monocytes subtypes (**Fig. 2C**). Indeed, 98 of these genes were directly associated with NF-κB-related pathways, highlighting its potential role in shaping distinct monocyte functions (**Fig. 2D**). Analysis of the temporal dynamics revealed that, at low and mid doses, the proportion of non-classic monocytes increased at both the EOI and 24h relative to the Pre time point (**Fig. 2E**). Given that non-classic monocytes are linked to tissue repair and inflammation resolution, their elevation, along with the enrichment of NF-κB signaling-related genes in monocytes subtypes, reflects the anti-inflammatory effects of ILB-202 (*29*). Notably, the high dose group exhibited a different trend from the low and mid dose groups showing a slight decrease, which could be attributed to the higher ILB-202 concentration leading to mild side effects (**Fig. 2E**).

Therefore, we conducted a cell-cell interaction analysis to investigate how ILB-202 shapes intercellular signaling. Despite the relatively low number of time-dependent transcript-level DEGs, this network-based approach revealed marked changes in intercellular signaling pathways over time (**Fig. 2F-G**). Compared with the Pre time point, interactions associated with dampening inflammatory responses became more prominent at later time points. In particular, TGFβ signaling pathway (TGFB1-TGFBR1_TGFBR2 interaction), known for its anti-inflammatory and immunosuppressive properties, was consistently significant at both EOI (prob: 0.0034) and 24h time point (prob: 0.005) (**Fig. 2F-G**) (*30*). Visfatin signaling (NAMPT–INSR interaction) became significant at 24 h (prob: 0.001) and is noteworthy given the multifaceted role of NAMPT in regulating NF-κB and inflammatory processes (*31, 32*).

Conversely, the TNF signaling pathways (TNF-TNFRSF1A and TNF-TNFRSF1B interactions) showed reduced activity at later time points, signifying a transition toward the resolution of inflammation (**Fig. 2F-G**) (*33*). Additionally, the probabilities and p-values for the CCL signaling pathway (CCL5-CCR1 interaction) and the CXCL signaling pathway (CXCL8-CXCR2 interaction) were insignificant at EOI and 24h, further supporting a progressive reduction in pro-inflammatory signals in the presence of ILB-202 (**Fig. 2F-G**) (*34, 35*). Taken together, these findings suggest that ILB-202 promotes anti-inflammatory activity by modulating intercellular signaling pathways associated with immune regulation.

## 4. DISCUSSION

In the phase 1 clinical trial, we evaluated the safety, tolerability, and preliminary pharmacodynamic effects of the systemic administration of ILB-202, an allogeneic-engineered EV derived from HEK293 cells designed to selectively inhibit hyperactivated NF-κB in inflamed cells while minimizing broad immunosuppression. Our findings demonstrate that ILB-202 is safe and well-tolerated across all tested dose levels in healthy volunteers, with no serious or dose-limiting toxicities. Notably, only minor changes in immune cell profiles were identified, such as a mild decrease in NK cell counts and a single case of grade 1 neutropenia in the high-dose cohort. Additionally, the absence of significant alterations in laboratory parameters, vital signs, or cytokine levels indicates a favorable safety profile, devoid of infusion-related reactions or cytokine release syndrome.

Although broad transcriptional changes were minimal in healthy volunteers, scRNA-seq revealed subtle but distinct alterations in signaling pathways associated with NF-κB, accompanied by a downregulation of key pro-inflammatory signaling networks. The time-dependent modulation of pathways involving TGFβ, visfatin (NAMPT–INSR), and TNF signaling indicates a potential shift toward a reduced inflammatory state. Overall, these pharmacodynamic effects highlight the anti-inflammatory effects of ILB-202 and its capacity for selective immune regulation without broadly suppressing gene expression in healthy tissues. In contrast, small-molecule NF-κB inhibitors (e.g., IKK inhibitors or proteasome inhibitors) often cause significant immunosuppressive side effects by suppressing basal immune activity and targeting upstream components of the NF-κB pathway that regulate multiple cellular processes (*17–19*).

Notably, this is the first clinical trial to investigate systemically administered, allogeneic-engineered EVs, marking a significant milestone in EV-based therapeutics. Previous EV-based therapies, such as mesenchymal stem cells derived exosomes tested for conditions such as COVID-19-induced ARDS (*10, 36–38*) and chronic inflammatory diseases (*39*)—relied on general anti-inflammatory properties for immune modulation and tissue repair. Although effective at reducing inflammation, they lack the pathway-specific targeting necessary to address NF-κB–driven hyperinflammation. Therefore we believe that the standardized cargo-loading process of ILB-202 for selective NF-κB inhibition provides a more robust and precise therapeutic approach. Additionally, the successful demonstration of both safety and pharmacodynamic activity not only validates the concept of engineered EVs as a therapeutic modality but also paves the way for future clinical development in diverse disease contexts. These results are particularly promising for the application of engineered EV therapeutics in inflammatory and autoimmune disorders, where the precise modulation of immune pathways is essential.

Consequently, future studies should investigate ILB-202 in conditions characterized by excessive or dysregulated NF-κB activation to further confirm its therapeutic potential, safety, and efficacy in clinically relevant populations. In conclusion, the results of this phase I trial highlighted the target mechanism of action and favorable safety profile of ILB-202, demonstrating the its potential as a next-generation immunomodulator for the treating of inflammatory and autoimmune disorders.

## 5. Author Contributions

S.H., H.C., Y.S.: Formal analysis; data curation; visualization, writing - original draft; writing – review & editing; D.H., S.-H.A., K.C., S.R., C.P.: study design, data analysis, review of manuscript; H.Y.G. & C.C.: study design, writing and review of manuscript

## 6. Data Availability Statement

The data that support the findings of this study are available on request from the corresponding author. The data are not publicly available due to privacy or ethical restrictions.

## Notes

This study was supported by a grant from the Drug Development Program through the Korea Health Industry Development Institute (HI20C0170), funded by the Ministry of Health and Welfare, Republic of Korea.C.C. is the founder and shareholder of ILIAS Biologics, Inc.; H.C. is the shareholder and ex-employee of ILIAS Biologics, Inc.; S.H, D.H, S-H.A., K.C., S.R. and C.P. are employee of ILIAS Biologics, Inc. The remaining authors declare no competing interests.

### Competing Interest Statement

C.C. is the founder and shareholder of ILIAS Biologics, Inc.; H.C. is the shareholder and ex-employee of ILIAS Biologics, Inc.; S.H, D.H, S-H.A., K.C., S.R. and C.P. are employee of ILIAS Biologics, Inc. The remaining authors declare no competing interests.

### Clinical Trial

NCT05843799

### Funding Statement

This study was supported by a grant from the Drug Development Program through the Korea Health Industry Development Institute (HI20C0170), funded by the Ministry of Health and Welfare, Republic of Korea.

### Author Declarations

Ethics committee of Bellberry Human Research Ethics Committee gave ethical approval for this work.

