## Supplementary materials for "Safety and anti-inflammatory effects of ILB-202, an engineered extracellular vesicles for NF-κB inhibition: A double-blind, randomized, placebo-controlled phase 1 trial"

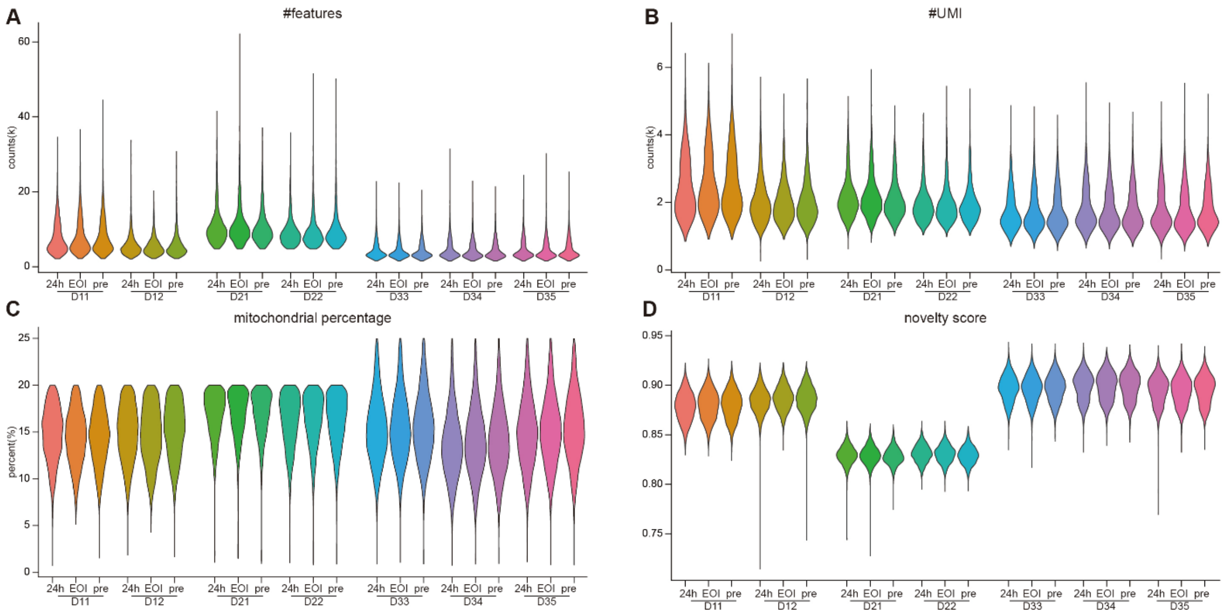

**Supplementary Figure 1.** Summary of quality control metrics for signle-cell RNA sequencing.

**A-D,** Violin plots representing QC metrics across samples: (A) number of detected genes, (B) unique molecular identifier (UMI) counts, (C) mitochondrial RNA percentage, and (D) novelty scores, calculated as the log10-transformed number of detected genes divided by the log10-transformed number of UMI counts per cells.

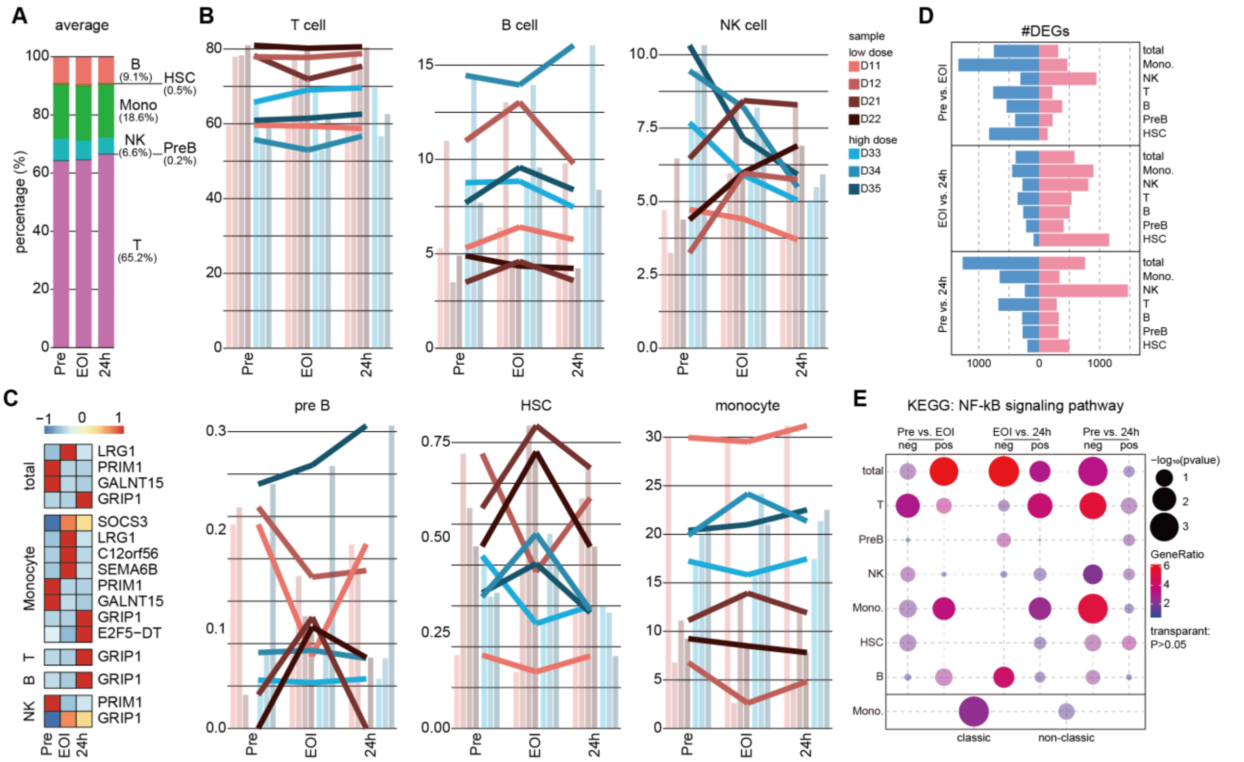

**Supplementary Figure 2.** Dynamic patterns of cell type proportions and differentially expressed genes (DEGs) across time points.

**A**, Bar plot showing the proportion of each cell type across the combined samples at each time point (Pre, EOI, and 24h). The plot represents the average distribution of cell types over time. **B**, Bar plots showing the proportion of each cell type at each time point for individual samples, with overlaid lines representing the trend in proportions for each individual over time. **C**, Heatmap showing the expression levels of genes identified as differentially expressed (DEGs) (**Fig. 2B**), including total DEGs and those specific to each cell type. **D**, Bar plot showing the number of DEGs identified under relaxed criteria ($|\log_2(\text{fold change})|>0$ and raw p-value $<0.05$) for each comparison: Pre vs. EOI, EOI vs. 24h, and Pre vs. 24h. Blue bars represent the number of downregulated genes, while pink bars indicate the number of upregulated genes. Compared to Figure 2B, the relaxed threshold results in a higher number of DEGs. **E**, Dot plot showing the results of gene set enrichment analysis (GSEA) for the KEGG NF-κB signaling pathway using DEGs identified in panel B. The bottom row represents the enrichment analysis performed separately for DEGs in classic and non-classic monocytes. Dot size corresponds to $-\log_{10}$(p-value), while dot color indicates the gene ratio. Transparent dots represent pathways with p-value ≥ 0.05.

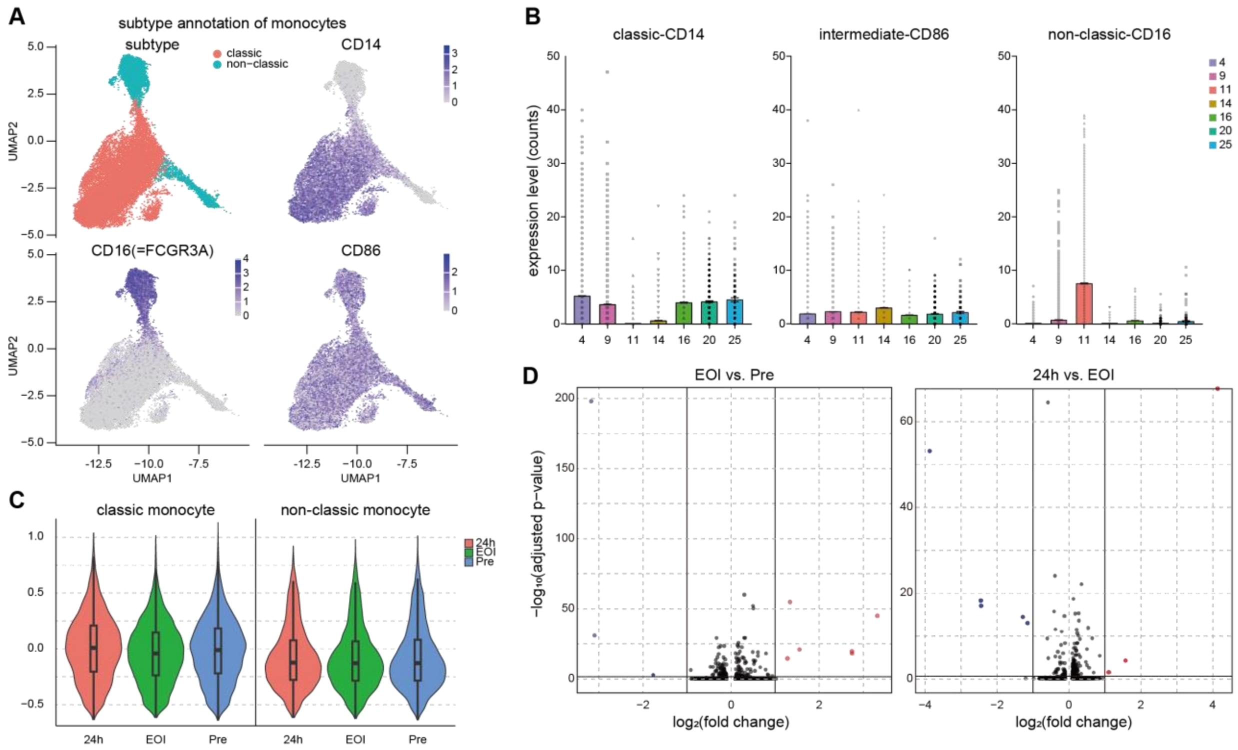

**Supplementary Figure 3.** Monocyte subtype annotation.

**A**, UMAP of monocytes distinguished by subtype (classic, non-classic), and the expression levels of marker genes. CD14, a marker for classic monocytes, shows high expression in the classic subtype, CD16 (FCGR3A), a marker for non-classic monocytes, is highly expressed in the non-classic subtype; and CD86, a marker for intermediate monocytes, displays variable expression among the subtypes. **B**, Bar plots showing the expression levels of marker genes for monocyte subtypes: CD14 (classic), CD86 (intermediate), and CD16 (non-classic) across monocyte clusters. These marker gene expression patterns were used to classify each cluster into classic, or non-classic monocyte subtypes. **C**, Violin plots showing the module scores for classic monocyte markers and non-classic monocyte markers at each time point (Pre, EOI, and 24h). The scores represent the aggregated expression of marker genes, highlighting the temporal changes in marker activity within monocyte subtypes. **D**, Volcano plots showing DEGs in monocytes across time points. The left plot compares Pre vs. EOI, and the right plot compares EOI vs. 24h. Genes with increased expression in the control group are shown on the left side of each plot, while those with increased expression in the comparison group are displayed on the right side.

**Supplementary Table 1. Full inclusion and exclusion criteria**

| Inclusion criteria |
| --- |
| 1. Must have given written informed consent before any study-related activities are carried out and must be able to understand the full nature and purpose of the trial, including possible risks and adverse effects.<br>2. Aged 18 to 55 years of age (inclusive) at the time of consent.<br>3. A body mass index (BMI) of ≥ 18.0 and ≤ 30.0 kg/$m^2$ at Screening.<br>4. Medically healthy male or female volunteers, without clinically significant abnormalities (in the investigator's opinion) at Screening and Day 1 (pre-dose), including:<br>a. Physical examination without (in the investigator's opinion) any clinically relevant findings;<br>b. Systolic blood pressure in the range of 90 to 160 mmHg and diastolic blood pressure in the range of 50 to 95 mmHg after at least 5 minutes in supine/ semi-supine position;<br>c. Pulse rate in the range of 45 to 100 bpm after at least 5 minutes rest in supine/semi-supine position;<br>d. Body temperature (tympanic), between 35.5°C and 37.7°C;<br>e. No clinically significant findings (in the investigator's opinion) in serum chemistry, hematology, coagulation, and urinalysis tests.<br>5. Conventional 12-lead ECG recording in triplicate (the mean of triplicate measurements will be used to determine eligibility at Screening, Day -1 and pre-dose on Day 1) consistent with normal cardiac conduction and function, including:<br>a. Normal sinus rhythm with HR between 45 and 100 bpm, inclusive;<br>b. QT interval corrected using the Fridericia method (QTcF) between 350 to 450 msec for male volunteers and 350 to 470 msec for female volunteers, inclusive;<br>c. QRS duration of < 120 msec;<br>d. PR interval of 120-220 msec; |

e. Electrocardiogram morphology consistent with healthy cardiac ventricular conduction and normal rhythm, and with measurement of the QT interval.

6. Female volunteers:

   a. Must be of non-child-bearing potential (i.e., surgically-sterilized [hysterectomy, bilateral salpingectomy, bilateral oophorectomy at least 6 weeks before the Screening visit]) or post-menopausal (where post-menopausal is defined as no menses for 12 months without an alternative medical cause, and a follicle-stimulating hormone [FSH] level >40 IU/L at the Screening visit), or

   b. If of child-bearing potential, must agree not to donate ova, not to attempt to become pregnant and, if engaging in sexual intercourse with a male partner, must have been using an acceptable forms of highly effective contraception (refer to Appendix AC) for at least 1 month prior to Screening (that period will extend to 3 months for oral contraceptive use), and agree to continue to use an acceptable form of contraception from the time of signing the consent form until at least 90 days after the dose of the IP.

   c. The male partner of a female study subject with child-bearing potential must use a condom from the time of the female subject signing the consent form until at least 90 days after the dose of the IP. *Note: this will not apply to male volunteers who are infertile and do not produce sperm (i.e., surgically-sterilized or biologically infertile, subject to the conditions detailed in Appendix AC*).

   d. All women of child-bearing potential must return a negative serum pregnancy test result at Screening and a negative urine pregnancy test before receipt of IP.

7. Male volunteers:

   a. Must agree to abstain from sperm donation and if engaging in sexual intercourse with a female of child-bearing potential must agree to use a condom and be willing to comply with the contraceptive requirements of the study, from the time of signing the consent form at Screening until at least 90 days after the dose of the IP. *Note: this will not apply to male volunteers who are infertile and do not produce sperm (i.e., surgically-sterilized or biologically infertile, subject to the conditions detailed in Appendix AC*).

8. Must have adequate bilateral venous access to allow for dose infusion and repeated blood sampling.

9. Must be able to communicate well with the PI and site personnel and agree to comply with all study procedures and requirements.

Exclusion criteria

1. Have clinical signs of active infection (inclusive of positive COVID-19 test result, where applicable) and/or a tympanic temperature of > 38.0°C at the time of screening. *Note: Study entry may be deferred at the discretion of the PI.*
2. Any condition requiring the regular use of medication.
3. Any condition which, while not requiring regular use of medication, is likely to require intermittent (as required) therapeutic intervention (e.g., migraine, seizure disorder, asthma).
4. History or evidence of any clinically significant condition (including and not limited to the gastrointestinal, renal, hepatic, cardiovascular, respiratory, endocrine, hematological, psychiatric or neurological systems) and/or other major disease or malignancy, as determined by the PI.
5. Family history of short or long QT syndrome.
6. History of risk factors for torsade de pointes or the diagnosis.
7. History of malignant disease in the last 10 years (excluding surgically resected skin squamous cell or basal cell carcinoma).
8. Presence or evidence of recent sunburn, scar tissue, tattoo (more than 25% of body area), open sore, or branding that, in the opinion of the investigator, would interfere with interpretation of infusion site reaction assessments.

9. History of drug allergies (including to any excipients) including a history of anaphylactic reaction.
10. History of or positive results of serology screening for hepatitis B (hepatitis B surface antigen [HBsAg]), hepatitis C (anti-hepatitis C virus [HCV] antibodies) or HIV (HIV antibodies type 1 and 2).
11. History of drug or alcohol abuse within 12 months prior to Day -1 or evidence of such abuse on laboratory assays at Screening and Day -1. Alcohol abuse defined as > 10 units of alcohol consumed per week (1 unit = ½ pint beer, 25 mL of 40% spirit or a 125 mL glass of wine).
12. History of alcohol consumption in the 48 hours prior to Day -1.
13. Clinically significant abnormalities in the screening physical examination, vital signs, ECG or clinical laboratory tests, as determined by the PI. Repeat testing may be performed at the PI's discretion.
14. ALT/AST > 1.5 ULN, total bilirubin > 1.5 x ULN and/or creatinine clearance < 80mL/min (calculated using Cockcroft & Gault formula).
15. Individuals who have smoked cigarettes or tobacco within 1 month prior to Day -1 or have a positive urine cotinine result at Screening or Day -1.
16. Exposure to any prescription medications (small molecules) or, topically or systemically administered over the counter drugs, dietary supplements, or herbal remedies, (with the exception of contraceptives for female participants of child-bearing potential, and the occasional use of paracetamol [no more than 2 g per day on no more than 3 days in one week]), within 14 days or 5 half-lives (if known), (whichever is longer) prior to the start of IP administration on Day 1.
17. Received treatment with immune-suppressive or immune-modulative medication (including topical, systemic and inhalant corticosteroids) or have received immunoglobulins and/or blood products within 90 days prior to the administration of the first dose of IP).
18. Have received treatment with or taken non-steroidal anti-inflammatory drugs (NSAIDs) within 14 days or 5 half-lives (if known), (whichever is longer) prior to the start of IP administration on Day 1.

19. Exposure to biologics within 6 months prior to Day -1. Influenza and COVID-19 vaccines administered > 14 days prior to the start of IP administration on Day 1 will be allowed.
20. Participation in another clinical trial of an investigational small molecule (or medical device) within 30 days (or 5 half-lives of the drug, whichever is longer) prior to the start of IP administration on Day 1 (or within 6 months prior to the start of IP administration on Day 1 if the investigational drug was a biologic).
21. Female volunteers who are pregnant or lactating.
22. Unable to refrain from strenuous activity for 3 days prior to Day -1 and for the duration of the study.
23. Major surgery within 90 days prior to Day -1 or anticipated surgery during the study.
24. Blood or plasma donation of > 100 mL during the 3 months prior to Day -1.
25. Unlikely to comply with the clinical study protocol: e.g., uncooperative attitude, unwilling to refrain from smoking during the study period, unwilling to reside in the study unit for the duration of the study, inability to return for follow-up visits, or improbability of completing the study.
26. Any other condition, which in the opinion of the PI precludes the volunteer's participation in the clinical study.

**Supplementary Table 2. Restricted Medications**

| **Product Category** | **Timeframes in which product is prohibited:** |
|---|---|
| Any prescription medications (small molecules) or topically or systemically administered over-the-counter drugs, dietary supplements, or herbal remedies.<br><br>The occasional use of paracetamol (up to a maximum of 2g per day on no more than 3 days in 1 week) will be allowed for the treatment of headache or other pain, subject to prior approval from the investigator.<br><br>*Note: Hormonal contraceptives for female participants of child-bearing potential are allowed.* | Within 14 days or 5 half-lives (if known), (whichever is longer) prior to the start of IP administration on Day 1, and at any time during the study period. |
| Immunosuppressive or immunomodulative medication (including topical, systemic and inhalant corticosteroids); or immunoglobulins and/or blood products. | Within 90 days prior to the start of IP administration on Day 1, and at any time during the study period. |
| Non-steroidal anti-inflammatory drugs (NSAIDs). | Within 14 days or 5 half-lives (if known), (whichever is longer) prior to the start of IP administration on Day 1, and at any time during the study period. |
| Biologics<br><br>*Note: Influenza and COVID-19 vaccines administered > 14 days prior to administration of the IP on Day 1 will be allowed.* | Within 6 months prior to the start of IP administration on Day 1, and at any time during the study period. |

**Supplementary Table 3. Summary of treatment-emergent adverse events by system organ class and preferred term (Safety analysis set)**

| SOC/PT | Pooled Placebo (n=6) n (%) | ILB-202 Low dose (n=4) n (%) | ILB-202 Meidum dose (n=4) n (%) | ILB-202 High dose (n=4) n (%) | Pooled ILB-202 (n=12) n (%) | Overall (n=18) n (%) |
|---|---|---|---|---|---|---|
| **Number of subjects with at least one TEAE** | **2 (33.3)** | **3 (75.0)** | **3 (75.0)** | **4 (100.0)** | **10 (83.3)** | **12 (66.7)** |
| **Blood and lymphatic System Disorders** | - | - | - | 1 (25.0) | 1 (8.3) | 1 (5.6) |
| Neutropenia | - | - | - | 1 (25.0) | 1 (8.3) | 1 (5.6) |
| **Gastrointestinal Disorders** | - | 2 (50.0) | - | - | 2 (16.7) | 2(11.1) |
| Abdominal pain upper | - | 1 (25.0) | - | - | 1 (8.3) | 1 (5.6) |
| Diarrhoea | - | 1 (25.0) | - | - | 1 (8.3) | 1 (5.6) |
| **General disorders and administration site conditions** | 2 (33.3) | - | 2 (50.0) | 3 (75.0) | 5 (41.7) | 7 (38.9) |
| Catheter site pain | - | - | - | 1 (25.0) | 1 (8.3) | 1 (5.6) |
| Infusion site pain | 2 (33.3) | | 1 (25.0) | 1 (25.0) | 2 (16.7) | 4 (22.2) |
| Infusion site discomfort | - | - | - | 1 (25.0) | 1 (8.3) | 1 (5.6) |
| Infusion site erythema | - | - | 1 (25.0) | - | 1 (8.3) | 1 (5.6) |
| Infusion site pruritus | - | - | 1 (25.0) | - | 1 (8.3) | 1 (5.6) |
| Non-cardiac chest pain | - | - | - | 1 (25.0) | 1 (8.3) | 1 (5.6) |
| **Nervous System Disorders** | - | - | 1 (25.0) | 1 (25.0) | 2 (16.7) | 2 (11.1) |
| Dizziness | - | - | - | 1 (25.0) | 1 (8.3) | 1 (5.6) |
| Headache | - | - | 1 (25.0) | - | 1 (8.3) | 1 (5.6) |
| **Respiratory, thoracic and mediastinal disorders** | - | 1 (25.0) | - | - | 1 (8.3) | 1 (5.6) |
| Oropharyngeal pain | - | 1 (25.0) | - | - | 1 (8.3) | 1 (5.6) |

Abbreviations: TEAE = treatment-emergent adverse event; n = Number of subjects reporting at least one TEAE in that category.

| |
|---|
| Notes: Treatment-emergent AEs are defined as AEs that occurred or worsened from the first administration of study drug up to 8 (+3) days after study drug administration. If a subject experienced the same AE multiple times, this was only to be counted once per System Organ Class and Preferred Term for the purpose of counting the number of subjects experiencing that AE. Adverse events that were experienced multiple times with different relationships/severities were only to be counted once per System Organ Class and Preferred Term for the worst-case relationship with study drug/severity. |

**Supplementary Table 4. Summary of immune cell marker results (immunogenicity analysis set)**

| | ILB-202 Low dose (n=4) | | ILB-202 Medium dose (n=4) | | LB-202 High dose (n=4) | | Pooled ILB-202 (n=12) | | Pooled Placebo (n=6) | | Overall (n=18) | |
|---|---|---|---|---|---|---|---|---|---|---|---|---|
| | Mean (SD) | Mean Δ Baseline (SD) | Mean (SD) | Mean Δ Baseline (SD) | Mean (SD) | Mean Δ Baseline (SD) | Mean (SD) | Mean Δ Baseline (SD) | Mean (SD) | Mean Δ Baseline (SD) | Mean (SD) | Mean Δ Baseline (SD) |
| **CD4+ (%grandparent)** | | | | | | | | | | | | |
| Baseline | 37.6 (3.0) | - | 43.8 (12.2) | - | 32.5 (6.4) | - | 38.0 (8.8) | - | 44.4 (4.8) | - | 40.1 (8.2) | - |
| 24 hr Post-Dose | 37.1 (2.4) | -0.5 (3.3) | 45.0 (8.0) | 1.3 (5.2) | 35.6 (8.9) | 3.1 (4.2) | 39.2 (7.7) | 1.3 (4.2) | 44.2 (2.6) | -0.2 (2.6) | 40.9 (6.8) | 0.8 (3.7) |
| 48 hr Post-Dose | 36.5 (2.4) | -1.1 (2.9) | 44.9 (7.1) | 1.1 (6.9) | 35.7 (7.9) | 3.2 (3.5) | 39.0 (7.2) | 1.1 (4.7) | 43.7 (2.5) | -0.7 (3.0) | 40.6 (6.3) | 0.5 (4.2) |
| **CD8+ (%grandparent)** | | | | | | | | | | | | |
| Baseline | 31.3 (2.5) | - | 26.0 (4.8) | - | 30.2 (5.7) | - | 29.2 (4.7) | - | 25.8 (4.9) | - | 28.1 (4.9) | - |
| 24 hr Post-Dose | 31.4 (2.5) | 0.1 (2.1) | 28.1 (5.6) | 2.1 (1.2) | 30.2 (7.4) | 0.0 (2.1) | 29.9 (5.2) | 0.7 (2.0) | 26.25 (4.9) | 0.4 (1.5) | 28.7 (5.3) | 0.6 (1.8) |
| 48 hr Post-Dose | 32.1 (2.9) | 0.8 (2.1) | 27.7 (4.7) | 1.7 (0.9) | 30.0 (6.3) | -0.2 (0.7) | 29.9 (4.8) | 0.7 (1.5) | 25.4 (4.9) | -0.4 (1.4) | 28.4 (5.1) | 0.4 (1.5) |
| **CD19+ (%grandparent)** | | | | | | | | | | | | |
| Baseline | 14.6 (4.8) | - | 12.7 (7.9) | - | 11.0 (3.4) | - | 12.8 (5.4) | - | 12.9 (3.4) | - | 12.8 (4.7) | - |
| 24 hr Post-Dose | 14.2 (4.8) | -0.4 (1.2) | 12.2 (7.0) | -0.5 (2.2) | 13.0 (6.4) | 2.0 (3.1) | 13.1 (5.6) | 0.4 (2.4) | 12.7 (3.2) | -0.2 (1.2) | 13.0 (4.8) | 0.2 (2.1) |
| 48 hr Post-Dose | 14.2 (4.1) | -0.4 (2.0) | 10.5 (3.5) | -2.1 (4.9) | 12.7 (5.2) | 1.8 (2.3) | 12.5 (4.2) | -0.3 (3.4) | 13.5 (3.2) | 0.6 (1.1) | 12.8 (3.8) | 0.0 (2.9) |
| **CD16/56+ (%grandparent)** | | | | | | | | | | | | |

| | | | | | | | | | | | | |
|---|---|---|---|---|---|---|---|---|---|---|---|---|
| Baseline | 7.4 (3.0) | - | 9.1 (7.2) | - | 13.6 (6.1) | - | 10.0 (5.8) | - | 8.3 (3.8) | - | 9.4 (5.2) | |
| 24 hr Post-Dose | 7.7 (2.8) | 0.3 (1.8) | 6.4 (1.7) | -2.7 (7.3) | 9.1 (3.9) | -4.5 (2.3) | 7.70 (2.9) | -2.3 (4.6) | 8.0 (2.9) | -0.2 (3.4) | 7.8 (2.8) | -1.6 (4.2) |
| 48 hr Post-Dose | 7.7 (2.7) | 0.4 (2.01) | 8.2 (2.7) | -0.9 (6.8) | 10.4 (3.8) | -3.2 (5.7) | 8.8 (3.1) | -1.2 (5.0) | 8.2 (2.3) | -0.1 (2.9) | 8.6 (2.8) | -0.9 (4.4) |
| Abbreviations: SD = standard deviation; Δ baseline = change from baseline.<br>Notes: %Grandparent was defined as the percentage of events (cells) within each CD4+, CD8+, CD19+ and CD16/56+ cell population relative to the CD45+ population | | | | | | | | | | | | |
